# Availability of clinical trial individual patient data in the BMJ before and after adoption of a stringent data-sharing policy, compared with recent rates at other major medical journals

**DOI:** 10.64898/2026.05.15.26353284

**Authors:** Alison Avenell, Dorothy V.M. Bishop

**Affiliations:** Aberdeen Centre for Evaluation, University of Aberdeen, Foresterhill, Scotland; Department of Experimental Psychology, University of Oxford, UK

**Keywords:** open data, clinical trials, medical journals

## Abstract

**Background:** In 2024, the BMJ updated its data-sharing policy for clinical trials, requiring open deposit of deidentified individual participant data (IPD) before publication. We considered whether data-sharing increased after introduction of the new policy, and compared rates of data-sharing in 2025 with those of other leading medical journals.

**Method:** All data-sharing statements were downloaded from BMJ trials published in 2023 (submitted pre-updated policy) and 2025 (submitted post-updated policy). Data for 2025 were gathered for trials in five comparison medical journals that had less stringent data-sharing requirements. Data-sharing statements were coded to specify whether IPD were immediately available, and if not, why. We did not check quality of IPD or reproducibility of results.

**Results:** Openly available IPD for BMJ trials increased from 0/32 in 2023 to 19/33 (58%) after the updated policy; seven articles gave repository links that did not yield any data. In comparison journals, most authors provided data-sharing statements, but rates of open IPD varied from 0% to 5.6%.

**Conclusions:** There was a substantial increase in open sharing of IPD after introduction of the new BMJ policy compared to a prior period, and compared to other journals in 2025.

## Introduction

Provision of deidentified individual participant (IPD) data allows assessment of the reproducibility and robustness of those data, helps assure data longevity, provides accountability and transparency, and makes best use of participants’, researchers’ and funders’ investment (Munafò et al., 2017; Nosek et al., 2022).

In 2016, the International Committee of Medical Journal Editors (ICMJE) put forward a proposal for promoting data sharing for clinical trials, which was published simultaneously in the Annals of Internal Medicine, British Medical Journal, JAMA (Journal of the American Medical Association), NEJM (New England Journal of Medicine) The Lancet, and eight other medical journals (Taichman et al, 2016). This stated that researchers had an ethical obligation to "to share with others the deidentified individual-patient data (IPD) underlying the results presented in the article (including tables, figures, and appendices or supplementary material) no later than 6 months after publication".

In response to feedback, a modified recommendation was published in 2017 (Taichman et al., 2017), which required clinical trials to have a data-sharing statement, but fell short of mandating open IPD, while continuing to "envision a global research community in which sharing deidentified data becomes the norm." Two new journals (Bulletin of the World Health Organisation, and Korean Journal of Medical Science) were signatories to the 2017 recommendation - but three of the original signatories (Canadian Medical Association Journal, Chinese Medical Journal and Nederlands Tijdschrift voor Geneeskunde) were absent; it was unclear whether they found the new recommendation too onerous, too weak, or whether there were other reasons for not signing.

Danchev et al (2021) looked at trials published in JAMA, Lancet and NEJM after they had adopted the ICMJE policy, and found that whereas 334/487 articles had a statement about data-sharing, only two IPD datasets were deidentified and openly available. Bergeat et al (2022) considered data-sharing for randomised controlled trials (RCTs) published in ten leading surgical journals, comparing 65 studies published before the policy was implemented and 65 published after. A data-sharing statement was provided for 11 of 65 RCTs after the policy versus none before the policy, but actual sharing was rare - IPD were available for two studies prior to the policy and two after the policy. Hamilton et al (2023) performed a systematic review of articles that evaluated data-sharing of medical studies in the period 2016-2021, and estimated that the proportion stating they would openly share IPD was 8%, whereas the proportion who actually shared was 2%.

In 2015, the BMJ went beyond the ICMJE recommendations in introducing a requirement that clinical trials share IPD on request (Loder & Groves, 2015). In 2024, the BMJ strengthened its data-sharing policy to make sharing of IPD and code mandatory prior to publication for RCTs (Loder et al., 2024).

The value of data-sharing was subsequently reiterated in an editorial by the Editor-In-Chief (Abbasi, 2025), after serious questions were raised about the integrity of data underpinning a trial of stem cell injection to prevent heart failure after myocardial infarction, leading to the trial’s retraction from the BMJ (Attar et al., 2025). Scrutiny of the open peer review documents showed that IPD were deposited in an open repository only after two requests by the editor, and after peer review had been conducted. Abbasi stated "*One purpose of mandatory data sharing is that it might act as a deterrent. This would now seem to be a false hope. A reasonable conclusion from this episode is that transparency and data sharing are not enough to create trust. We’re therefore examining how to prevent a similar situation arising again. It’s clear that greater scrutiny of the underlying data is required before publication."*

This case piqued our interest in the extent to which fully open data from clinical trials can be achieved if a policy of mandatory data-sharing is taken seriously by editors, so that authors understand they cannot simply ignore it. Abbasi (2025) noted that BMJ had refused to publish papers where authors were unable or unwilling to share data. This provides an interesting natural experiment, allowing us to compare the extent of data-sharing for papers submitted before and after the introduction of the more stringent data-sharing policy in May 2024. For comparison, we assessed data-sharing at five other prominent journals that publish clinical RCTs (Annals of Internal Medicine, JAMA, Nature Medicine, New England Journal of Medicine, and The Lancet), four of whom had been signatories of the 2017 statement on data sharing, and all of which are current members of ICMJE and have policies regarding data-sharing or data-sharing statements (see Table 1). As well as considering rates of open data-sharing, we did an ad hoc qualitative analysis of the content of data-sharing statements, to gain some insight into reasons given for data availability restrictions when data are not shared.

**Table 1.**
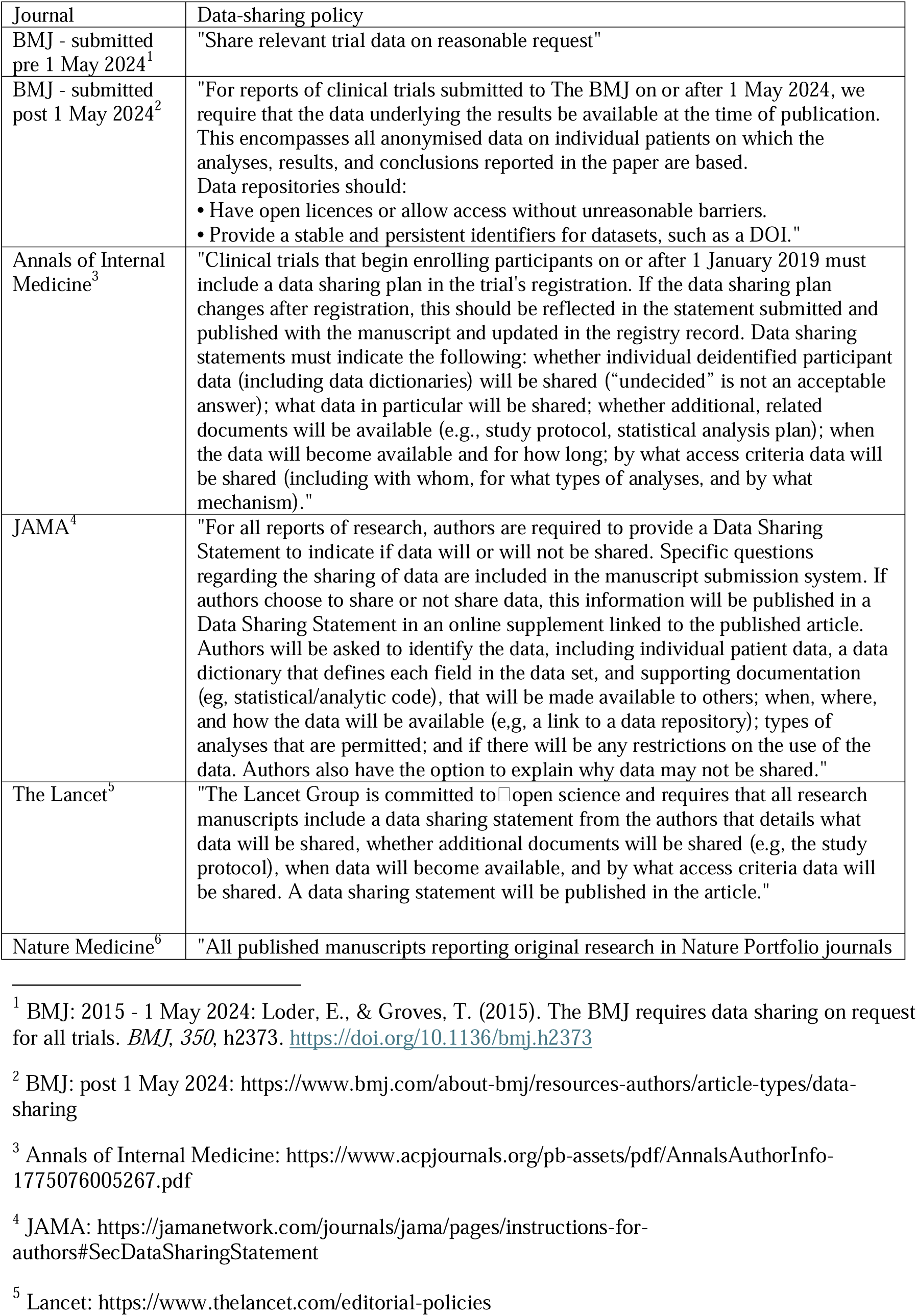
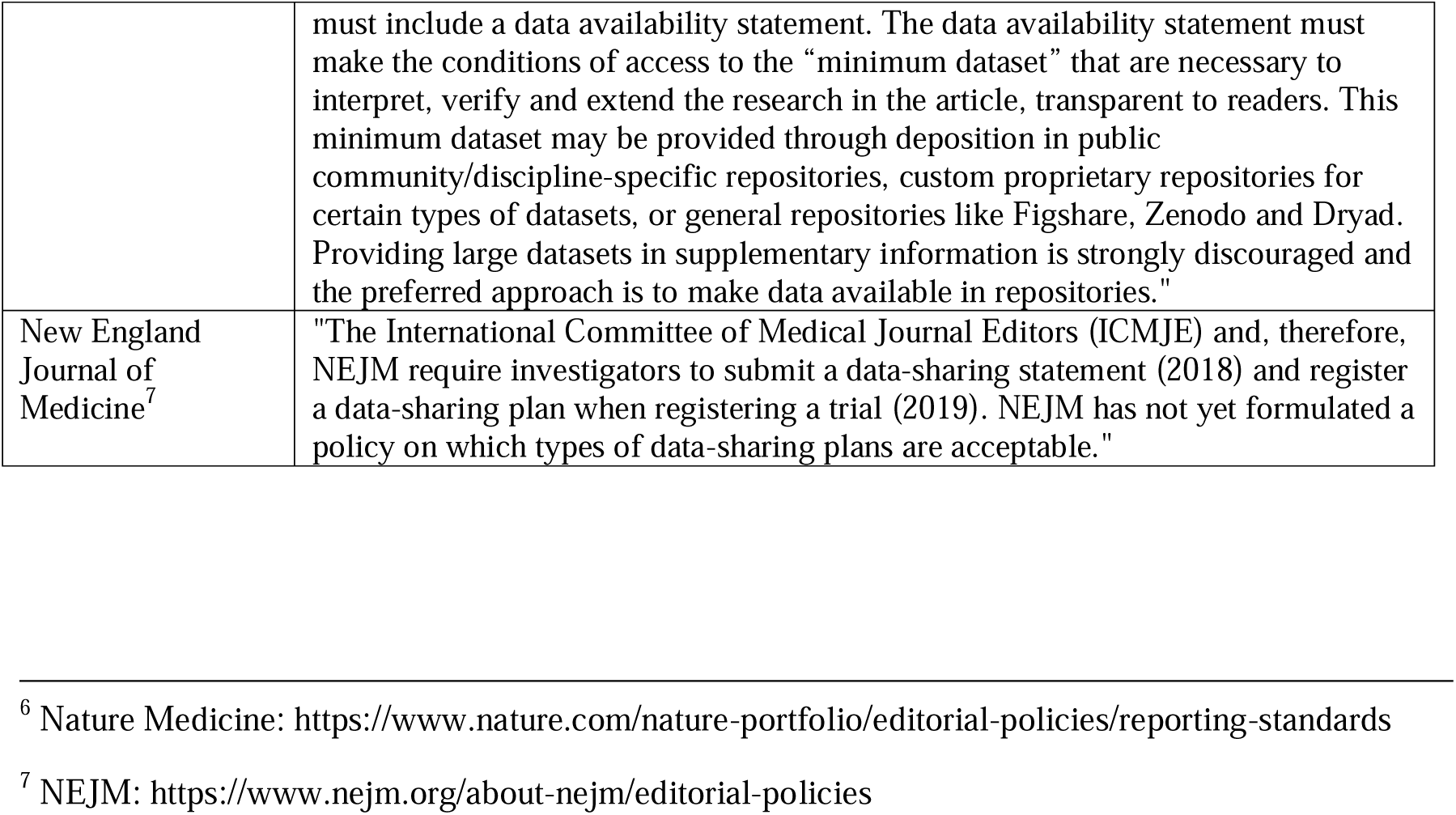
Data-sharing policies of six leading medical journals

Our aims were to examine:

1. How often clinical trial IPD are shared in other major medical journals that ask authors to provide data-sharing statements?
2. Whether mandating openly accessible IPD in the BMJ was associated with an increase in trials providing IPD?
3. Where IPD are not available, what conditions do authors specify to allow access?

## Methods

In addition to the BMJ, we selected the five highest impact factor general medical journals, with one only from each publisher (Annals of Internal Medicine, JAMA, Nature Medicine, NEJM, and The Lancet), all of which routinely publish RCTs. RCTs in each journal were identified by a search of the Dimensions database for articles with the keywords "trial" or "RCT" for the year of 2025. For the BMJ the same search was conducted for the period 1st January 2023 to 30^th^ April 2024, before the more stringent data-sharing policy was introduced (1st May 2024). We considered also searching the other journals for the earlier period, but they had had no change to data sharing policies.

The authors scrutinised the resulting list of articles to exclude review articles, meta-analyses, or other non-RCT articles, leading to the dataset summarised in Table 2.

**Table 2.**
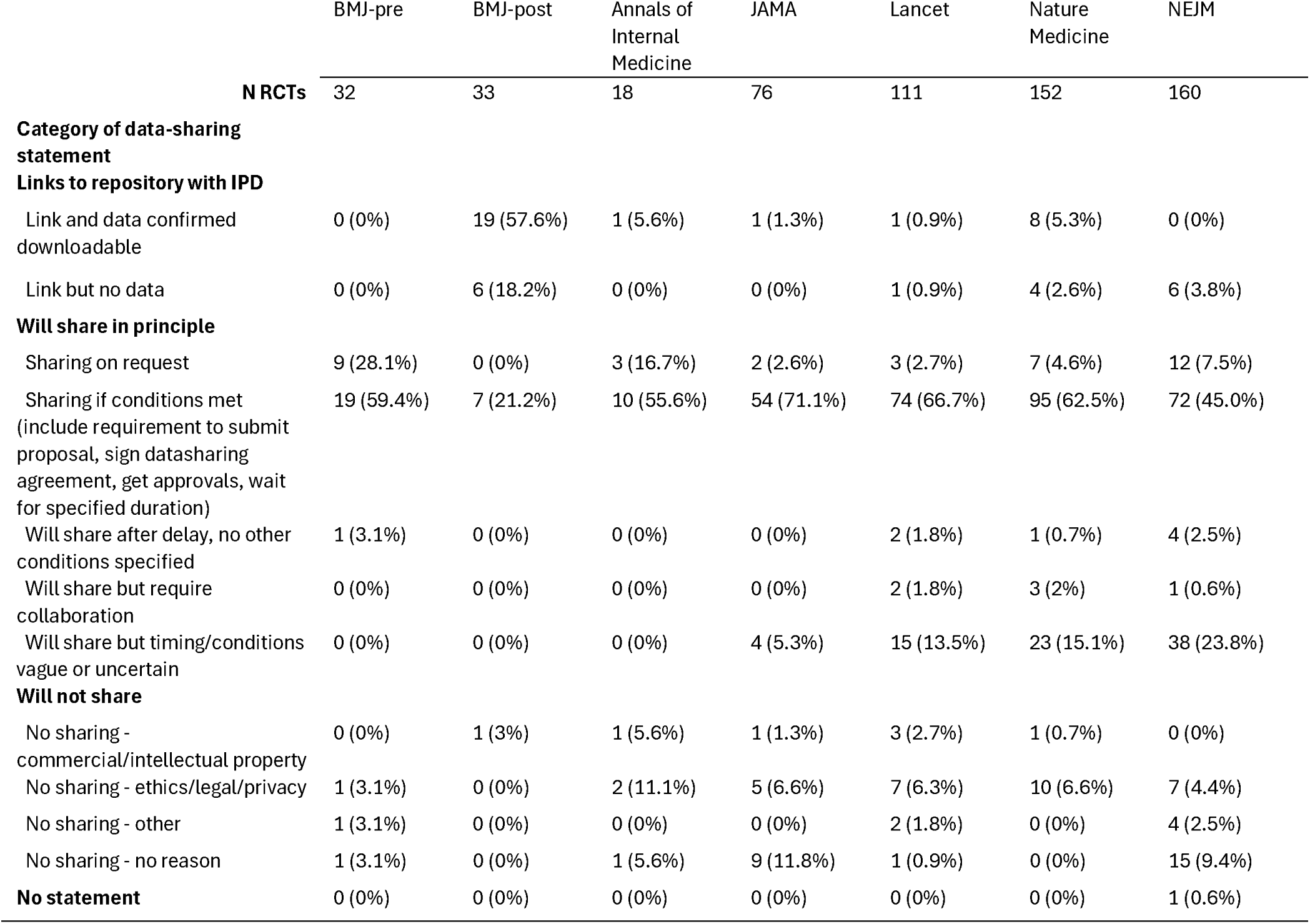
Numbers (%) of articles categorized according to data-sharing statements.

|  | BMJ-pre | BMJ-post | Annals of Internal Medicine | JAMA | Lancet | Nature Medicine | NEJM |
| --- | --- | --- | --- | --- | --- | --- | --- |
| **N RCTs** | 32 | 33 | 18 | 76 | 111 | 152 | 160 |
| **Category of data-sharing statement** |  |  |  |  |  |  |  |
| **Links to repository with IPD** |  |  |  |  |  |  |  |
| Link and data confirmed downloadable | 0 (0%) | 19 (57.6%) | 1 (5.6%) | 1 (1.3%) | 1 (0.9%) | 8 (5.3%) | 0 (0%) |
| Link but no data | 0 (0%) | 6 (18.2%) | 0 (0%) | 0 (0%) | 1 (0.9%) | 4 (2.6%) | 6 (3.8%) |
| **Will share in principle** |  |  |  |  |  |  |  |
| Sharing on request | 9 (28.1%) | 0 (0%) | 3 (16.7%) | 2 (2.6%) | 3 (2.7%) | 7 (4.6%) | 12 (7.5%) |
| Sharing if conditions met (include requirement to submit proposal, sign datasharing agreement, get approvals, wait for specified duration) | 19 (59.4%) | 7 (21.2%) | 10 (55.6%) | 54 (71.1%) | 74 (66.7%) | 95 (62.5%) | 72 (45.0%) |
| Will share after delay, no other conditions specified | 1 (3.1%) | 0 (0%) | 0 (0%) | 0 (0%) | 2 (1.8%) | 1 (0.7%) | 4 (2.5%) |
| Will share but require collaboration | 0 (0%) | 0 (0%) | 0 (0%) | 0 (0%) | 2 (1.8%) | 3 (2%) | 1 (0.6%) |
| Will share but timing/conditions vague or uncertain | 0 (0%) | 0 (0%) | 0 (0%) | 4 (5.3%) | 15 (13.5%) | 23 (15.1%) | 38 (23.8%) |
| **Will not share** |  |  |  |  |  |  |  |
| No sharing - commercial/intellectual property | 0 (0%) | 1 (3%) | 1 (5.6%) | 1 (1.3%) | 3 (2.7%) | 1 (0.7%) | 0 (0%) |
| No sharing - ethics/legal/privacy | 1 (3.1%) | 0 (0%) | 2 (11.1%) | 5 (6.6%) | 7 (6.3%) | 10 (6.6%) | 7 (4.4%) |
| No sharing - other | 1 (3.1%) | 0 (0%) | 0 (0%) | 0 (0%) | 2 (1.8%) | 0 (0%) | 4 (2.5%) |
| No sharing - no reason | 1 (3.1%) | 0 (0%) | 1 (5.6%) | 9 (11.8%) | 1 (0.9%) | 0 (0%) | 15 (9.4%) |
| **No statement** | 0 (0%) | 0 (0%) | 0 (0%) | 0 (0%) | 0 (0%) | 0 (0%) | 1 (0.6%) |

Our primary interest was in whether IPD were openly available. We found that it was necessary to check this to the point of locating the dataset, because there were cases where links were provided to a repository but data could not be found, or were inaccessible. We coded data as openly available if the only requirement for access was registering on the repository where the data were held, but not if other conditions had to be met. In a few cases, a subset of data was openly deposited; these articles were coded as having data available if deposited data were sufficient to repeat the analysis in principle. (An example would be a case where demographic data were not open, but data were provided for primary and secondary outcomes).

We did not attempt to reproduce findings reported in the articles, nor to evaluate the quality and integrity of deposited data.

For articles without open IPD, we coded data-sharing statements according to whether they indicated that sharing of data was possible (coded "maybe"), or explicitly stated that data would not be shared (coded "no"), and recorded whether a data-sharing platform was used. We also devised an ad hoc coding scheme to categorise statements in terms of whether there were time-based or other restrictions on data access, or if specific reasons were given to justify not sharing. One author coded all articles using this scheme. Its reliability was checked by two other researchers independently analysing 100 of the 584 articles.

All data-sharing statements are included in the spreadsheet documenting the articles and coding for this study (see data-sharing statement). In some cases these gave an explicit rationale for restricting data access, or gave conditions for sharing. Illustrative examples of these were extracted to provide a more qualitative perspective on data-sharing. These extracts are not intended to be representative or comprehensive, but rather to show the diversity of justifications researchers gave for their statements.

In response to reviewer suggestions, we conducted a supplementary analysis on additional information as follows:

a. The total sample size for each study. This information was usually taken from the Abstract (included in our main data workbook), but the full text was consulted for cases where it was missing from the Abstract.
b. The sponsor of the study. To obtain this information, we first identified the trial registry from the PubMed entry for each study (or from the original text in a few cases where it was missing) and then extracted information about the study Sponsor from the registries. Sponsors were categorised as either industry or non-industry.
c. For studies where IPD were available from a link provided by authors, the F-UJI data assessment tool (Devaraju & Huber, 2025) was used to evaluate adherence to FAIR principles. The authors of this tool caution that it is under development and cannot evaluate certain aspects of FAIR that depend on human judgement, but it can quantify those features that are assessable by automation.
d. For studies with a commercial sponsor that specified a data-sharing platform for data requests, we searched the data-sharing platform for the trial registry number to establish whether the study was registered on that platform.

## Results

### Reliability of the coding scheme for data sharing statements

For the basic 3-way categorisation for data being available (yes), possibly available (maybe), or not available (no), agreement was near perfect for all pairs of three raters (97%, 98% and 99%). For the "maybe" category, we initially attempted to distinguish statements that specified a delay period prior to proposals being considered for approval, and those that did not, but the language used in the data-sharing statements was often unclear, leading to poor agreement. We therefore merged these categories to give the broad category of "Sharing if conditions met". The pairwise agreement for the more granular codes shown in Table 2 was still imperfect at 81%, 86% and 84%, but we felt this was adequate to give a more qualitative impression of the relative frequency of different conditions that researchers specified for data-sharing. For the final categorisation of data-sharing, we used consensus codes for articles that had been multiply-coded, and the coding by one author (DB) for the remainder.

### Rates and conditions of data-sharing

Results are shown in Table 2.

Our first question was how often clinical trial IPD are shared in journals that ask authors to provide data-sharing statements. It is clear that if authors are asked to provide data-sharing statements, they do so, but these very seldom indicate that IPD will be shared without any restrictions.

There was a pronounced increase in the proportion of articles with open IPD in the BMJ since introduction of mandatory data-sharing policy in 2024: pre-policy BMJ data, 0/32 (0%) v post-policy 19/33 (57.6%). Rates of open IPD in BMJ prior to the policy are comparable to those seen in the comparison journals in 2025, which ranged from 0% to 5.6%.

There were seven BMJ trials published after the BMJ changed its policy where we could not access data, despite statements that data were available. Four trials had links to the Dryad repository. Correspondence with Dryad revealed that data from these trials had never been deposited. The BMJ was notified of missing data from trials on 28th February 2026. As of 11th May 2026 no revisions or notices had been posted by the BMJ for these trials, and data remained unavailable.

For the comparison journals, where a data-sharing statement was provided, it was relatively uncommon for authors to state they would share data "*on (reasonable) request*". A typical data-sharing statement would request that researchers submit a proposal for approval and require a data-sharing agreement to be signed if approval was given. Frequently, there were additional conditions to be met, concerning the time frame of requests (often only possible after a delay), the qualifications of the research team, and/or approvals from ethics committees. Some data-sharing statements were too vague to be useful.

Supplementary Table 1 shows data on total sample size for a study, and on sponsorship, with studies subdivided into the 29 which were open (i.e. link to a repository with minimal barriers to sharing IPD) versus the remainder. For three journals, BMJ, Annals of Internal Medicine and JAMA, rates of industry sponsorship were low, whereas for the Lancet, Nature Medicine and NEJM around half the studies were industry-sponsored. The sample is too small for statistical comparison, but there was no suggestion that submissions to BMJ were different before and after the new data-sharing policy, either in terms of sample size of studies, or commercial sponsorship.

For the 29 studies where authors provided a link to IPD data in a repository, results from the F-UJI data assessment tool for evaluating adherence to FAIR principles seemed largely a function of the specific repository. Datasets from Dryad, Dataverse and Scidb were rated as Advanced, those from Figshare, Zenodo, Mendeley, Handle, Fairdata, Open Science Framework, Synapse and Canadian Research Data Repository as Moderate.

For studies sponsored by pharmaceutical companies, data-sharing was often handled by specific data-sharing platforms, notably Vivli, ClinicalStudyDataRequest and YODA (see Supplementary Table 2). These platforms were set up to encourage data reuse, while controlling access to ensure responsible use. Requests for data are submitted via a standard form, and evaluated by an independent committee, and users are required to sign a Data Use Agreement. These were categorised as "Sharing if conditions met" in Table 2. A check of the trial registration number against these repositories found a match for 12/80 on Vivli, 1/10 on YODA, and 2/6 on ClinicalStudyDataRequest. Five of the matched Vivli records stated that a data package was available for download pending approval

### Illustrative reasons given for restricting data access

Extracts from data-sharing statements illustrate the range of reasons given for restricting data access. These examples are extracted from the spreadsheet with all data (DOI: 10.6084/m9.figshare.32270577).

L033: "*Gilead Sciences shares anonymised individual participant data upon request or as required by law or regulation with qualified external researchers based on submitted curriculum vitae and reflecting non-conflict of interest. The request proposal must also include a statistician. Approval of such requests is at Gilead Science’s discretion and is dependent on the nature of the request, the merit of the research proposed, the availability of the data, and the intended use of the data."*

N033: (In response to question When will data availability begin?) "*After the medicine and indication is approved by both the FDA and EMA*."

N061: "*Investigators who have invested time and effort into developing a trial or study should have a period of exclusivity in which to pursue their aims with the data, before key trial data are made available to other researchers."*

A018: "*The PHRI believes the dissemination of clinical research results is vital and sharing of data is important. PHRI prioritizes access to data analyses to researchers who have worked on the trial for a substantial duration, have played substantial roles, and have participated in raising the funds to conduct the trial. PHRI balances the length of the research study, and the intellectual and financial investments that made it possible with the need to allow wider access to the data collected. Data will be disclosed only upon request and approval of the proposed use of the data by a Review Committee. Data will be available to the journal for evaluation of reported analyses. Data requests from other non-POISE-3 investigators will not be considered until 5 years after the closeout of the trial*."

Another concern that was sometimes explicitly stated was that providing data would be time-consuming for the original research group:

M090:"*Data requests will be considered by the Sponsor and the University of Glasgow, which will take account of the scientific rationale, ethics, logistics and resource implications Requests for transfer of deidentified data (including source imaging scans) will be considered by the University of Glasgow, and, if approved, a collaboration agreement would be expected, taking account of any cost implications. Cost recovery would be expected on a not-for-profit basis."*

Trials sponsored by pharmaceutical companies often gave a generic data-sharing statement. This reflected a commitment to data-sharing, in the context of needing to protect commercial interests and adhere to ethical standards, as well as preventing use of the data for unmotivated data-dredging. This statement from a study sponsored by Merck & Co illustrates these points:

M113: "*MSD is committed to providing qualified scientific researchers access to anonymized data and clinical study reports from the company’s clinical trials for the purpose of conducting legitimate scientific research Feasible requests will be reviewed by a committee of MSD subject matter experts to assess the scientific validity of the request and the qualifications of the requesters. In line with data privacy legislation, submitters of approved requests must enter into a standard data-sharing agreement with MSD before data access is granted. Data will be made available for request after product approval in the United States and the European Union or after product development is discontinued*"

J038: The data-sharing statement provided a link to a repository, but a request for the data led access being denied after answering No to a question "Do you agree to offer the BedMed co-investigators the opportunity to participate in research projects and publications that originate from the shared BedMed data?"

## Discussion

### Data sharing higher when mandated

These results suggest that, despite frequent statements by journal editors, drug companies, and researchers about the value of open data, immediate sharing of IPD is strongly resisted by most researchers. The case of the BMJ is important in suggesting that open data can be achieved if it is a condition for publishing in the journal, but this policy needs to be enforced by editorial teams who check compliance. For the remaining journals, data sharing statements were almost always provided, but usually with restrictions, such as time delays, agreeing the analysis, qualifications of the requestors, ethical approval, and cost involved to the trial team. In a minority of cases (ranging from 0% to 11.8%) data-sharing statements simply said data would not be shared, without giving any reason. A similar proportion justified not sharing because of ethical, legal or commercial considerations

### Reasons for widespread resistance to data sharing

Smith and Roberts (2016) outlined seven reasons that were given for not sharing data: (i) others using the data without having gone to the trouble of collecting them; (ii) being scooped; (iii) conclusions may not replicate; (iv) an alternative analysis could challenge published results; (v) sharing data may reveal weaknesses in data management; (vi) concerns about patient confidentiality; and (vii) technical reasons. Smith and Roberts declared all of these to be invalid. In contrast, Longo and Drazen (2016) argued that uncontrolled data sharing would encourage "data parasites", i.e. "people who had nothing to do with the design and execution of the study but use another group’s data for their own ends, possibly stealing from the research productivity planned by the data gatherers, or even use the data to try to disprove what the original investigators had posited" (p. 276). The illustrative examples of data-sharing statements reflected these concerns (2016), especially inappropriate use of data, allowing others to take the credit for the work, or being scooped.

Smith and Roberts (2016) were dismissive of such concerns, particularly where they appeared to reflect a "data-hoarding" approach, or involved barriers being erected to prevent the researchers from close scrutiny of their data management or datasets. Importantly, patients endorse sharing of their data, providing it is anonymised (National Academies of Sciences, Engineering and Medicine, 2020). Nevertheless, we know from numerous studies that, unless IPD are deposited with a permanent DOI, they are unlikely to ever be made available to other researchers, regardless of what the data-sharing statement says (Hussey, 2025; Kotoulas et al., 2026). Quite simply, websites decay and research groups disband and become uncontactable (Vines et al., 2014). Data-sharing is less onerous when planned for at the start of a project: reconstructing a dataset after the study is completed can involve much more work.

It is important, nevertheless, to recognise that there can be valid reasons for concerns about data misuse, as illustrated by recent examples of AI generation of pointless papers (Maupin et al., 2025), or use of cherrypicked data for ideological rather than scientific ends (Bishop, 2016). These issues seem characteristic of large open cohort datasets with hundreds of variables that allow for formulaic AI-assisted analyses of subsets of variables to generate numerous articles (Spick et al., 2026). Data from RCTs are less likely to be attractive for those generating AI-assisted articles at scale, but with the growth of easily-accessible methods for scraping open data, the potential danger exists. The challenge for journals and researchers is how to protect data from misuse without losing the benefits of open data-sharing.

In apparent compliance with data sharing, some authors gave links to a data repository but then failed to deposit their data. This appears to be a common issue with articles that cite a Dryad DOI: Spick (2026) identified articles in Europe PMC that included a Dryad DOI and found that around 10% failed to resolve to deposited data. The reasons here are unclear but it is possible that some authors exploit the fact that Dryad will issue a "reserved DOI" prior to publication, which reassures editors and peer reviewers, but does not translate to an open dataset after publication.

### Use of data-sharing platforms

As shown in Supplementary Table 2, some studies stated that data could be requested via independent data-sharing platforms. Such platforms were discussed at length in a workshop on clinical trial data sharing run by the US National Academies of Sciences, Engineering and Medicine (2020). Platforms such as Vivli use independent review panels to scrutinise requests for data-sharing and require data-use agreements to ensure that those using data do so ethically. The benefits of such platforms are that they provide a transparent access route, overseen by an independent data committee, and check that data are usable, while preventing inappropriate use. They also have disadvantages, however. These platforms do not act as repositories for long-term storage of data, and most studies whose sponsors used these platforms were not linked to the platform, suggesting that data would only be prepared for sharing after a request was approved. Most data would never become available. In addition, the costs of data-sharing platforms may not be justified by their apparent benefits, and it is unclear who should bear those costs. Finally, they are unsuited for one of the main reasons for data-sharing, i.e. for third parties solely to check the accuracy and integrity of a deposited dataset. The conditions for access imposed by data-sharing platforms cause delays and bureaucratic burdens on those seeking access to data; it was noted in the NASEM report that institutions often are slow to approve data-sharing agreements, and may wrangle over precise wording. These barriers may be insuperable for those who wish simply to check the accuracy of a deposited dataset. Models such as as FreeBIRD (https://freebird.lshtm.ac.uk/) and ImmPort (https://www.immport.org/shared/home), which make deidentified data sets available to those who register on the site and accept their terms, would be preferable.

### Study limitations

This study was not registered. It was an exploratory observational study, prompted by the retraction of a trial in the BMJ with untrustworthy IPD, where it was not possible to specify the details of methods until we had staked out the territory by familiarising ourselves with the range of data-sharing statements used by researchers. The methods evolved as we progressed through the coding, and some attempts at categorisation were abandoned as unreliable. To counteract this weakness, we have provided the spreadsheets underlying the analysis, including the data-sharing statements and our coding, so that coding decisions can be evaluated by readers, and others can build on this work if they find it useful.

In terms of identifying relevant papers, a reviewer suggested that the PRESS search strategy reporting guidelines, with input from an information specialist, would have been useful (McGowan et al., 2016). Our failure to do so was the result of ignorance of the guidelines.

This study relies on natural observational data, so cannot prove the policy change caused the increase in data-sharing seen in the BMJ. Furthermore, the coding used in Table 2 was devised by a process of trial and error to settle on categories that were both reasonably reliable and provided useful information, but its reliability is not established in a new dataset. Using a standardised form to collect details, for example, as undertaken by the NEJM, would likely remove some of the ambiguities in description which challenged us in our coding, providing that all questions were answered. We hope that other researchers working in this area may find the taxonomy reflected in our rating scale useful, but it remains to be seen if it is valid, especially in subject areas beyond clinical trials.

We did not have the resources to extend our analysis to check for open code, nor to examine whether reported results could be reproduced from available IPD. Once data are available, there are also valid questions about the quality of data provided, which may depend on the data-sharing platform. Whether data are complete and uncleaned or the final, cleaned dataset may be very important, both for questions of reproducibility and research integrity, but also replicability and new research.

### The growing need to check trial data integrity

It is notable that very few of the conditional data-sharing statements or journal statements in our study showed any appreciation of the value and need for data-sharing to check the integrity of data and analyses. There is growing concern that the number of fraudulent clinical trials is increasing, and it has been argued that sharing of IPD can help detect these (Van Noorden, 2023; Brown & Carlisle, 2025). The editor of the journal Anaesthesia (Carlisle, 2021), showed that checking IPD trial data flagged 26% of trial submissions as "zombie" trials, as opposed to 1% detection with trial level data. In a further study requesting IPD at submission to the journal (Bramley 2025), 87% of trial authors agreed to provide IPD, and 9% of authors reported finding errors in their data. Provision of IPD to journals for data integrity checks might be more acceptable to authors, but this would not constitute open access. Currently, many pharmaceutical researchers refuse to share IPD until product approval has been obtained; this makes sense from a commercial point of view, but it also implies that the data are error-free; in practice, if there are errors in data, it would be preferable to find them prior to product approval in the interests of patient safety.

Prominent medical journals are not immune to fraudulent data. Two of the journals in our analysis (the Lancet and NEJM) published analyses based on fraudulent (non-trial) registry data from Surgisphere, and it was noted that identification of the issues was hindered by inability to check original data (Editors of the Lancet, 2020; Mehra et al., 2020). Our current analysis was motivated by findings of data irregularities in a trial published in the BMJ, which has since been retracted. It is unlikely that this trial would have been questioned if data had not been openly available. Quite apart from such misconduct, there is ample evidence that honest errors in data and analysis are common, and unless we can detect them, they will remain to corrupt published research. Openly available data, without the need for permissions, would allow much faster identification and correction or retraction of studies with data irregularities.

### Future directions

Our analysis suggests that mandating immediate access to individual patient data on trial publication can work so long as journal editorial staff check that open data exist, rather than relying on statements to that effect. At minimum, random journal checks are needed to confirm that data are available when the authors give a link to a repository.

It is 10 years since the original ICMJE statement. Although there has been some progress since then, the landscape of trial integrity issues has been changed by increasing numbers of fraudulent studies (van Noorden, 2023). Journals may be forced to rethink their policies on clinical trial data-sharing in order to retain public confidence in the trials that they publish. In our view, it is time for the ICMJE to take the lead to strengthen enforcement of trial data-sharing; otherwise their statements will end up paying lip service to the need to trust trial data.

## Data Availability

The data and code used to create Table 2 are deposited at Figshare, DOI: 10.6084/m9.figshare.32270577.

https://doi.org/10.6084/m9.figshare.32270577

## Acknowledgements

We are most grateful to Mark Bolland for his helpful discussions about this article, and for contributing to data coding.

## Funding

None.

## Conflicts of interest

None to declare.

## Ethical approval

Not applicable.

---

**Supplementary Table 1.** Trial sample size and sponsorship by open data category.

|  | BMJ-pre | BMJ-post | Annals.Int<br>Med | JAMA | Lancet | NatMed | NEJM |
| --- | --- | --- | --- | --- | --- | --- | --- |
| **Open data (N)** | - | 19 | 1 | 1 | 1 | 8 | - |
| Median sample size | - | 420 | 108 | 150 | 686 | 82 | - |
| Min sample size | - | 45 | 108 | 150 | 686 | 6 | - |
| Max sample size | - | 5420 | 108 | 150 | 686 | 741 | - |
| Prop. industry sponsor | - | 0.11 | 0 | 0 | 0 | 0.12 | - |
| **Not open data (N)** | 32 | 14 | 17 | 75 | 110 | 144 | 160 |
| Median sample size | 612 | 566 | 171 | 641 | 670 | 206 | 758 |
| Min sample size | 51 | 50 | 101 | 72 | 11 | 3 | 6 |
| Max sample size | 48282 | 2159 | 5810 | 131276 | 357716 | 274182 | 332428 |
| Prop. industry sponsor | 0.06 | 0.07 | 0.12 | 0.16 | 0.50 | 0.48 | 0.52 |

**Supplementary Table 2.**
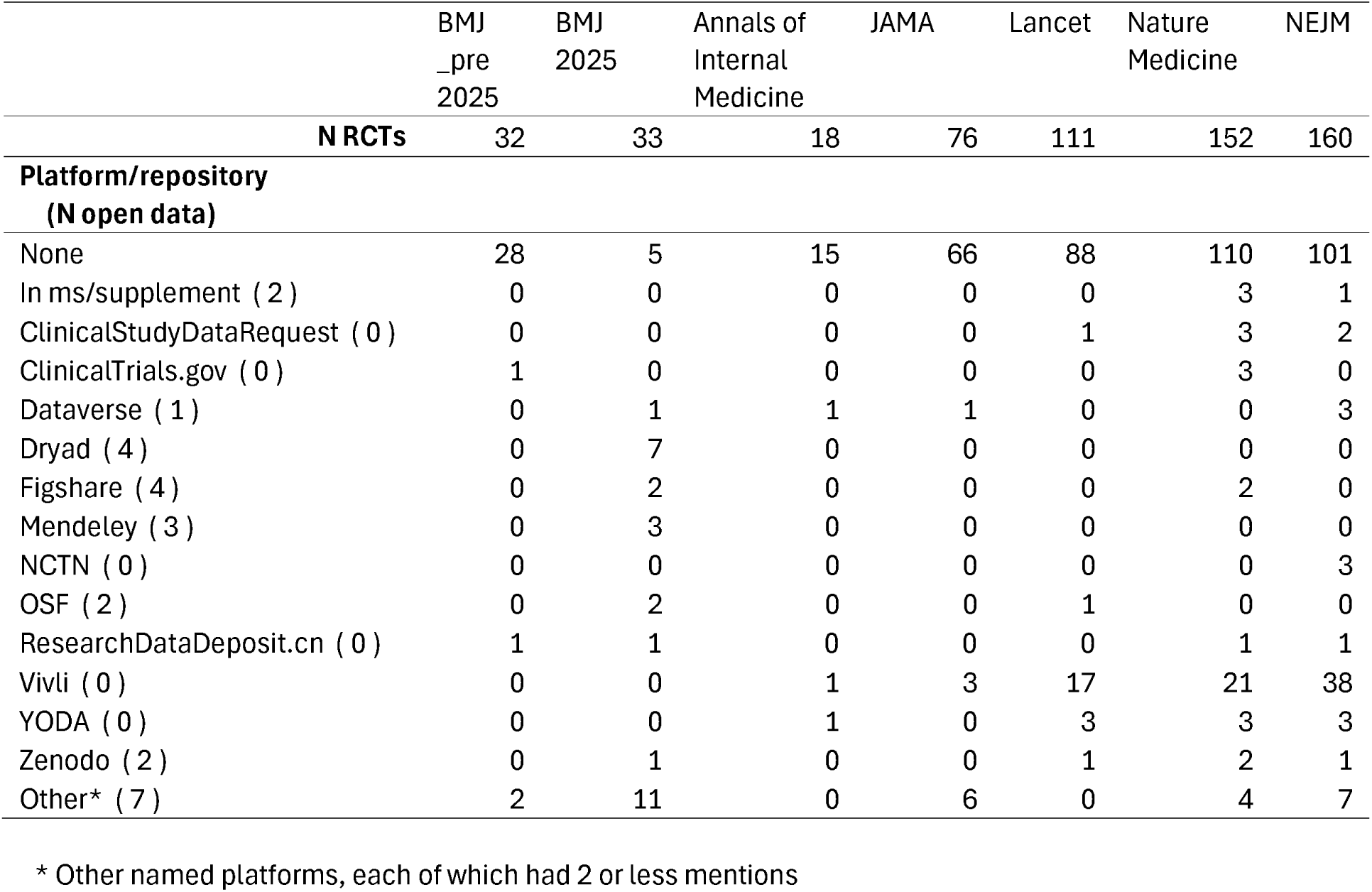
Data-sharing platforms/repositories mentioned by articles summarised in Table 2, with N with open data shown in brackets.

## Notes

### Competing Interest Statement

The authors have declared no competing interest.

### Summary of Updates

Some minor changes made to tables to correct with updated information. Title modified to clarify the method. We note that we did not check quality of IPD or reproducibility of results. We shortened the Introduction, without removing the context of the study. Limitations notes that the study was not registered. In a new supplementary Table (supplementary table 1) we looked at the following in relation to whether or not data were openly available: Sponsorship: We have added information about whether trials were sponsored by pharmaceutical companies. Sample size: We report on median and range for sample sizes of articles. We have added the following text to the Methods: "In response to reviewer suggestions, we conducted a supplementary analysis on additional information as follows: a) The total sample size for each study. This information was usually taken from the Abstract (now included in our main data workbook), but the full text was consulted for cases where it was missing from the Abstract. b) The sponsor of the study. To obtain this information, we first identified the trial registry from the PubMed entry for each study (or from the original text in a few cases where it was missing), and then extracted information about the study Sponsor from the registries. Sponsors were categorised as either industry or non-industry. c) For studies with a link to an online repository for IPD, the F-UJI data assessment tool (Devaraju & Huber, 2025) was used to evaluate adherence to FAIR principles. d) For studies with a commercial sponsor that specified a data-sharing platform for data requests, we searched the data-sharing platform for the trial registry number to establish whether the data had been deposited." We moved the selected data-sharing comments to the Results. We have strengthened our discussion of the need for more action towards the end of the discussion. We have made aims more specific: 1. How often clinical trial IPD are shared in other major medical journals that ask authors to provide data-sharing statements? 2. Whether mandating openly accessible IPD in the BMJ was associated with an increase in trials providing IPD? 3.Where IPD are not available, what conditions do authors specify to allow access? For those articles with a link to a repository, we used the F-UJI Automated FAIR Data Assessment Tool to evaluate adherence to FAIR principles, and we now report those results ( https://www.f-uji.net/index.php)

## References

Abbasi, K. (2025). The potential and limits of scrutiny in medical research. BMJ, 391, r2411. 10.1136/bmj.r2411

Attar, A., Mirhosseini, S. A., Mathur, A., Dowlut, S., Monabati, A., Kasaei, M., Abtahi, F., Kiwan, Y., Vosough, M., & Azarpira, N. (2025). Prevention of acute myocardial infarction induced heart failure by intracoronary infusion of mesenchymal stem cells: Phase 3 randomised clinical trial (PREVENT-TAHA8). BMJ, 391, e083382. 10.1136/bmj-2024-083382

Bergeat, D., Lombard, N., Gasmi, A., Le Floch, B., & Naudet, F. (2022). Data sharing and reanalyses among randomized clinical trials published in surgical journals before and after adoption of a data availability and reproducibility policy. JAMA Network Open, 5(6), e2215209. 10.1001/jamanetworkopen.2022.15209

Bishop, D. V. M. (2016). Open research practices: Unintended consequences and suggestions for averting them. (Commentary on the Peer Reviewers’ Openness Initiative). Royal Society Open Science, 3(4), 160109. 10.1098/rsos.160109

Bramley, P. (2025). Ask, and it shall be given you – individual patient data and code availability for randomised controlled trials submitted for publication. Anaesthesia, 80(2), 205–206. 10.1111/anae.16503

Brown, N. J. L., & Carlisle, J. B. (2025). Detecting inconsistencies and fraud in research data: Time for authors to share the data underlying their summary statistics as a matter of course. Anesthesia & Analgesia, 10.1213/ANE.0000000000007889. 10.1213/ANE.0000000000007889

Carlisle, J. B. (2021). False individual patient data and zombie randomised controlled trials submitted to Anaesthesia. Anaesthesia, 76(4), 472–479. 10.1111/anae.15263

Danchev, V., Min, Y., Borghi, J., Baiocchi, M., & Ioannidis, J. P. A. (2021). Evaluation of data sharing after implementation of the International Committee of Medical Journal Editors data sharing statement requirement. JAMA Network Open, *4*(1), e2033972. 10.1001/jamanetworkopen.2020.33972

Devaraju, A., & Huber, R. (2025). *F-UJI - An Automated FAIR Data Assessment Tool* (Version v3.5.1) [Computer software]. Zenodo. 10.5281/ZENODO.6361400

Editors of the Lancet. (2020). Learning from a retraction. The Lancet, 396(10257), 1056. 10.1016/S0140-6736(20)31958-9

Hamilton, D. G., Hong, K., Fraser, H., Rowhani-Farid, A., Fidler, F., & Page, M. J. (2023). Prevalence and predictors of data and code sharing in the medical and health sciences: Systematic review with meta-analysis of individual participant data. BMJ, 382, e075767. 10.1136/bmj-2023-075767

Hussey, I. (2025). Data is not available upon request. Meta-Psychology, 9. 10.15626/MP.2023.4008

Kotoulas, B., Blackwell, J., & Hardwicke, T. E. (2026). Has transparency improved in clinical psychology? A repeated cross-sectional study (2012, 2018, 2024) (Jn435_v1). *PsyArXiv*. https://osf.io/preprints/psyarxiv/jn435_v1/

Loder, E., & Groves, T. (2015). The BMJ requires data sharing on request for all trials. BMJ, 350, h2373. 10.1136/bmj.h2373

Loder, E., Macdonald, H., Bloom, T., & Abbasi, K. (2024). Mandatory data and code sharing for research published by the BMJ. BMJ, 384, q324. 10.1136/bmj.q324

Longo, D. L., & Drazen, J. M. (2016). Comment: Data Sharing. The New England Journal of Medicine, 374(3), 276–277. 10.1056/NEJMe1516564

Maupin, D., Spick, M., & Geifman, N. (2025). Safeguarding Open Science from exploitative practices. PLOS Medicine, 22(12), e1004851. 10.1371/journal.pmed.1004851

McGowan, J., Sampson, M., Salzwedel, D. M., Cogo, E., Foerster, V., & Lefebvre, C. (2016). PRESS Peer Review of Electronic Search Strategies: 2015 guideline statement. Journal of Clinical Epidemiology, 75, 40–46. 10.1016/j.jclinepi.2016.01.021

Mehra, M. R., Desai, S. S., Kuy, S., Henry, T. D., & Patel, A. N. (2020). Retraction: Cardiovascular disease, drug therapy, and mortality in Covid-19. N Engl J Med. DOI: 10.1056/NEJMoa2007621. | NEJM (Pt 2582). New England Journal of Medicine, *382*. https://www.nejm.org/doi/full/10.1056/NEJMc2021225

Munafò, M. R., Nosek, B. A., Bishop, D. V. M., Button, K. S., Chambers, C. D., Percie du Sert, N., Simonsohn, U., Wagenmakers, E.-J., Ware, J. J., & Ioannidis, J. P. A. (2017). A manifesto for reproducible science. Nature Human Behaviour, 1(1021) 10.1038/s41562-016-0021

National Academies of Sciences, Engineering, and Medicine (2020). Reflections on Sharing Clinical Trial Data: Challenges and a Way Forward: Proceedings of a Workshop (C. Shore, J. Hinners, E. Khandekar, & T. Wizemann, Eds). National Academies Press (US). http://www.ncbi.nlm.nih.gov/books/NBK559433/

Nosek, B. A., Hardwicke, T. E., Moshontz, H., Allard, A., Corker, K. S., Dreber, A., Fidler, F., Hilgard, J., Struhl, M. K., Nuijten, M. B., Rohrer, J. M., Romero, F., Scheel, A. M., Scherer, L. D., Schönbrodt, F. D., & Vazire, S. (2022). Replicability, robustness, and reproducibility in psychological science. Annual Review of Psychology, 73(Volume 73, 2022), 719–748. 10.1146/annurev-psych-020821-114157

Smith, R., & Roberts, I. (2016). Time for sharing data to become routine: The seven excuses for not doing so are all invalid [version 1; peer review: 2 approved, 1 approved with reservations]. F1000Research, *5*, 781. 10.12688/f1000research.8422.1

Spick, M. (2026). Detection arbitrage: The Dryad data deposition dodge. Octopus. 10.57874/ayce-8140

Spick, M., Onoja, A., Harrison, C., Stender, S., Byrne, J., & Geifman, N. (2026). Quantifying new threats to health and biomedical literature integrity from rapidly scaled publications and problematic research. Journal of Clinical Epidemiology, 193, 112203. 10.1016/j.jclinepi.2026.112203

Taichman, D. B., Backus, J., Baethge, C., Bauchner, H., Leeuw, P. W. de, Drazen, J. M., Fletcher, J., Frizelle, F. A., Groves, T., Haileamlak, A., James, A., Laine, C., Peiperl, L., Pinborg, A., Sahni, P., & Wu, S. (2016). Sharing clinical trial data: A proposal from the International Committee of Medical Journal Editors. PLOS Medicine, *13*(1), e1001950. 10.1371/journal.pmed.1001950

Taichman, D. B., Sahni, P., Pinborg, A., Peiperl, L., Laine, C., James, A., Hong, S.-T., Haileamlak, A., Gollogly, L., Godlee, F., Frizelle, F. A., Florenzano, F., Drazen, J. M., Bauchner, H., Baethge, C., & Backus, J. (2017). Data sharing statements for clinical trials: A requirement of the International Committee of Medical Journal Editors. JAMA, *317*(24), 2491–2492. 10.1001/jama.2017.6514

Van Noorden, R. (2023). Medicine is plagued by untrustworthy clinical trials. How many studies are faked or flawed? Nature, 619(7970), 454–458. 10.1038/d41586-023-02299-w

Vines, T. H., Albert, A. Y. K., Andrew, R. L., Débarre, F., Bock, D. G., Franklin, M. T., Gilbert, K. J., Moore, J.-S., Renaut, S., & Rennison, D. J. (2014). The availability of research data declines rapidly with article age. Current Biology, 24(1), 94–97. 10.1016/j.cub.2013.11.014

